# Denoising diffusion model for increased performance of detecting structural heart disease

**DOI:** 10.1101/2024.11.21.24317662

**Authors:** Christopher D. Streiffer, Michael G. Levin, Walter R. Witschey, Emeka C. Anyanwu

## Abstract

Recent advancements in generative artificial intelligence have shown promise in producing realistic images from complex data distributions. We developed a denoising diffusion probabilistic model trained on the CheXchoNet dataset, encoding the joint distribution of demographic data and echocardiogram measurements. We generated a synthetic dataset skewed towards younger patients with a higher prevalence of structural left ventricle disease. A diagnostic deep learning model trained on the synthetic dataset performed comparably to one trained on real data producing an AUROC=0.75(95%CI 0.72-0.77), with similar performance on an internal dataset. Combining real data with positive samples from the synthetic data improved diagnostic accuracy producing an AUROC=0.80(95%CI 0.78-0.82). Subgroup analysis showed the largest performance improvement across younger patients. These results suggest diffusion models can increase diagnostic accuracy and fine-tune models for specific populations.

## 1 Introduction

Generative artificial intelligence has undergone significant advancements, particularly in the domain of image generation. Generative adversarial networks (GANs) [1, 2], variational autoencoders (VAEs) [3], and, more recently, Diffusion Models [4–6] have demonstrated the ability to generate high-quality images by sampling from complex data distributions.

These methodologies have been further extended to enable conditional image generation, producing images based on a provided input signal [7–16]. Notable examples of this include DALL-E [12, 17] and Stable Diffusion [6] which have achieved impressive results producing images based on user provided text. Within the medical domain, generative models have been practically applied to tasks such as image-to-image generation [18], image denoising [19], and the generation of synthetic datasets [7–10, 20–26]. Synthetic datasets offer significant promise to the medical field as they have been shown to accurately recreate existing data distributions across many domains including radiology [7, 8, 21, 27, 28], dermatology [7, 29], and histopathology [7, 25] and can be utilized to improve diagnostic accuracy and generalizability of machine learning models [7, 21, 24, 26, 27, 29].

Within the field of medical imaging, machine learning models have demonstrated significant diagnostic potential across many different tasks and imaging modalities [30–34]. Convolutional neural networks (CNNs) have been used to diagnose conditions such as diabetic retinopathy [35], classify skin cancer [31], and detect pancreatic cancer on non-contrast computed tomography (CT) [32]. The application of machine learning to chest X-rays (CXRs) alone has been profound with models being able to estimate cardiovascular risk [36], detect type-2 diabetes [37], predict ejection fraction [38], and screen for structural heart disease [39]. These models have not only replicated the diagnostic capabilities of radiologists [40] but have also uncovered patterns within images that are imperceptible to the human eye [41]. The integration of such models into clinical practice holds the potential to address the growing shortage of board-certified radiologists in comparison to the increasing demand for imaging studies and create new screening modalities that can be accessed from anywhere with an internet connection.

Despite these advancements, models often fail to generalize to new datasets that differ from their training data [42–44]. This struggle to make a clinical impact is often attributed to dataset biases [45], differences in patient populations [46, 47], and model overfitting on provided training data. This is further exacerbated by the scarcity of medical data and institutional barriers to data sharing [43, 48]. Generative models present a promising approach for creating synthetic data by enabling the conditional generation of images based on features that matter in clinical contexts, such as patient demographics and disease characteristics. For example, synthetic data can be generated to augment the representation of underrepresented groups defined by factors such as age, race, and sex [7, 20, 49]. By tailoring these synthetic datasets to reflect diverse patient populations, generative models can help mitigate biases in the original data, thereby enhancing diagnostic accuracy and improving the generalizability of machine learning models [7, 20, 49].

This work builds on the recent advancements in deep learning, specifically the model developed by Bhave et al. [39] for identifying severe left ventricular hypertrophy (SLVH) and dilated left ventricle (DLV) using CXRs. The authors of that study provided a dataset [39, 50, 51] linking CXRs with transthoracic echocardiogram (TTE) measurements, including interventricular septal thickness at end-diastole (IVSd), left ventricular internal diameter at end-diastole (LVIDd), and left ventricular posterior wall distance at end-diastole (LVPWd). In this work, we aim to train a conditional diffusion model that learns the joint distribution of demographic data and continuous echocardiogram measurements to generate a high-quality synthetic dataset. We demonstrate that a diagnostic deep learning model trained on the synthetic dataset achieves similar performance to a model trained on the real dataset, and that a model trained on a combination of real and synthetic data improves overall diagnostic accuracy. We show that the models achieve similar performance across a cohort of patients from the University of Pennsylvania Health System (UPHS). Finally, we show that increasing the representation of specific age groups within the synthetic dataset substantially improves diagnostic accuracy across that population.

## 2 Results

### 2.1 Experiment Overview

Our experimental approach (Figure 1) consisted of training a denoising diffusion probabilistic model (DDPM) on a subset of the CheXchoNet dataset, then using this model to generate a synthetic dataset with a differing distribution from the base dataset. To evaluate the effectiveness of the synthetic dataset, we replicated Bhave et al.’s [39] methodology of training a deep learning model to detect the presence of severe left ventricular hypertrophy (SLVH) and dilated left ventricle (DLV). Within the paper, the terms synthetic and generated are used interchangeably when describing the dataset produced by the diffusion model.

**Fig. 1.**
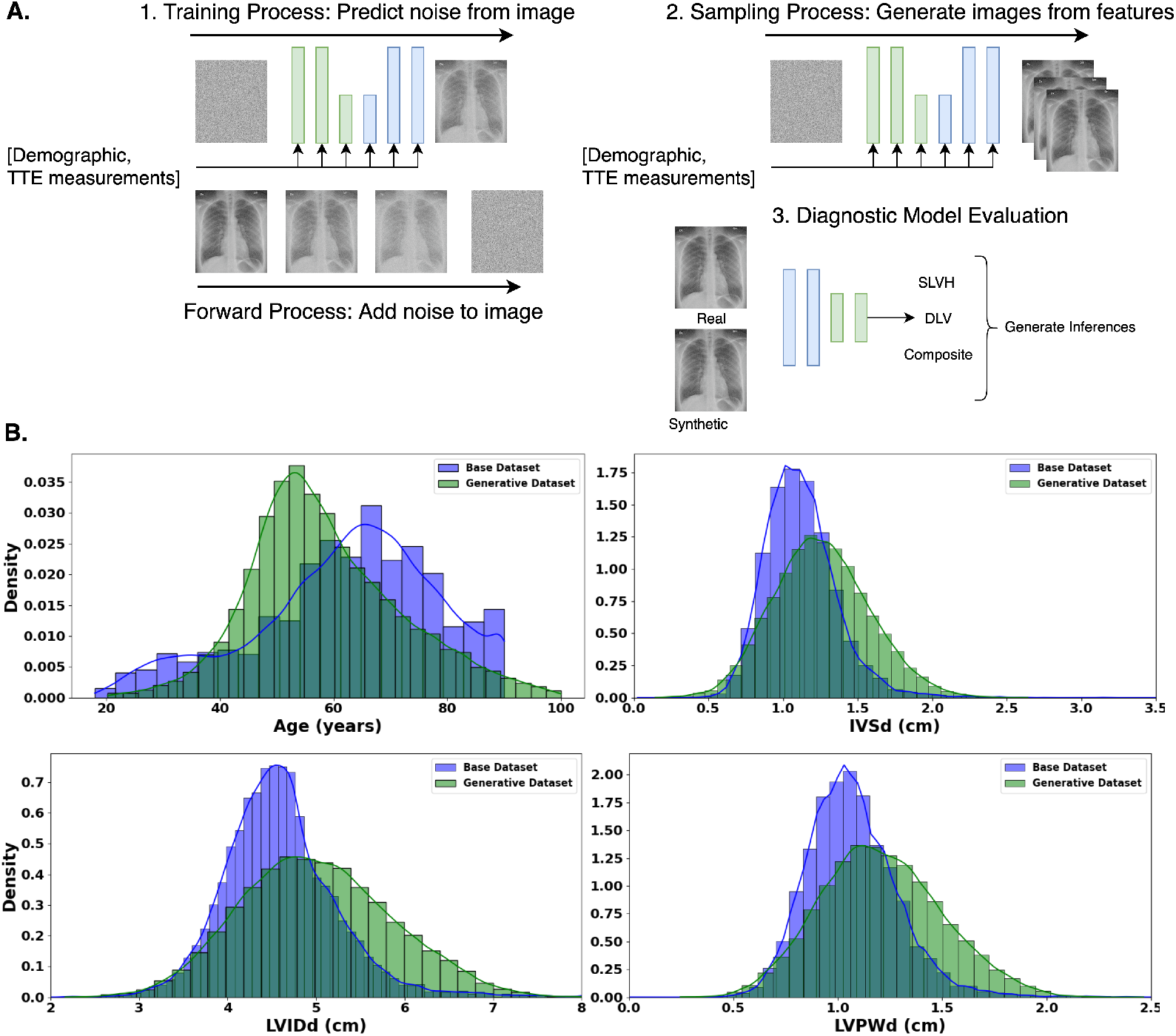
Overview of methodology consisting of diffusion model training, synthetic dataset generation, and diagnostic model evaluation. **A**. An overview of our methodology which consists of: **1**. Training the DDPM by first adding gaussian noise to the CXRs, then estimating the added noise conditioned on the input features of demographic data and echocardiogram measurements. **2**. Generating a synthetic dataset using the trained diffusion model by sampling from a specified distribution of gaussian noise and input features, then iteratively denoising the gaussian input conditional on the features to produce the output images. **3**. Training a deep learning model to predict the labels of SLVH, DLV, and composite on different partitions of real and synthetic data. **B**. Base (blue) and Generative (green) dataset distributions for age, IVSDd, LVIDd, and LVPWd.

We analyzed the performance of the model across four data partitions which included: a) the base dataset (Base), b) the generated dataset (Gen), c) a combination of the base and generated dataset (Base+Gen), and d) the base dataset combined with positive samples from the generated dataset (Base+Gen(Pos)). To prevent data leakage, we split the CheXchoNet dataset into training, validation, and testing partitions randomized by patient. The training dataset was used to train both the diffusion and diagnostic inference models, with the validation dataset used for model selection, and the testing dataset reserved for the final model evaluations.

### 2.2 Base Dataset vs Generated Dataset

The CheXchoNet dataset consisted of 64,277 Chest X-Rays and corresponding echocardiogram measurements taken from 22,220 unique patients. The mean age of the dataset was 62.3 (sd=16.0) with 37.8% of the patients being under the age of 60. Women were more accounted for in the dataset consisting of 56.6% of patients. The dataset had prevalence of SLVH and DLV of 8.6% and 6.0%, respectively. The dataset had echocardiogram measurements of IVSd 1.12 cm (sd=0.27), LVIDd 4.60 cm (sd=0.67), and LVPWd 1.07 cm (sd=.23).

The generated dataset was sampled to have a greater number of positive samples across a younger patient demographic. The mean age of the generated dataset was 58.6 (sd=13.4) with 57.5% of the samples being under the age of 60. Women accounted for 49.5% of samples. The dataset had prevalence of SLVH and DLV of 38.8% and 25.5%, respectively. The dataset had echocardiogram measurements of IVSd 1.26 cm (sd=0.32), LVIDd 5.01 cm (sd=0.86), and LVPWd 1.21 cm (sd=.29). We focused on this demographic because the original dataset had the highest proportion of negative samples across this age group. Further, accurately detecting structural heart disease at a younger age offers the benefits of earlier intervention and potentially greater long-term impact on patient outcomes.

We isolated positive samples from the generated dataset to create a more focused, combined dataset. The mean age of this positive sample dataset was 55.5 (sd=10.8), with patients below the age of 60 accounting for 70.7% of the total samples. The dataset had echocardiogram measurements of IVSd 1.44 cm (sd=0.321), LVIDd 5.48 cm (sd=0.82), and LVPWd 1.35 cm (sd=.29). Women accounted for 59.5% of samples. This gender imbalance occurred because echocardiogram measurements were uniformly sampled from a standard normal distribution, but the criteria for assigning SLVH and DLV labels are lower for women, resulting in their higher representation in these categories.

### 2.3 Conditional Diffusion Results

The performance of the diffusion model was evaluated through metrics that measured both the quality and diversity of the generated dataset, as well as the diagnostic accuracy in detecting structural heart disease. To assess image quality, the generated dataset was compared against the training and test datasets using the Frechet Inception Distance (FID) and Inception Score (IS). The FID metric measures the distance between two distributions and focuses more on image quality with the best possible score being 0, while the IS metric uses a pre-trained Inception [52] model to classify images into different categories which measures quality and diversity.

Our diffusion model produced an FID score of 9.62 compared to the training dataset and 13.5 compared to the testing dataset. For reference, the training dataset compared to the testing dataset produced an FID score of 4.07, which indicates that the synthetic dataset produces images that are similar to the base distribution. Our diffusion model produced an IS of 5.83 which was similar to the IS for the training and testing datasets of 5.71 and 5.82, respectively, indicating good quality and diversity of generated images. We have provided examples of generated images compared to real images that have been cross-matched based on age, sex, and TTE measurements (Figure 2). As can be observed, the generated images closely resembled the cross-matched images from the training and testing datasets. We have provided further details on the demographics of these images, as well as the results from the diagnostic models in Supplementary Table 3.

**Fig. 2.**
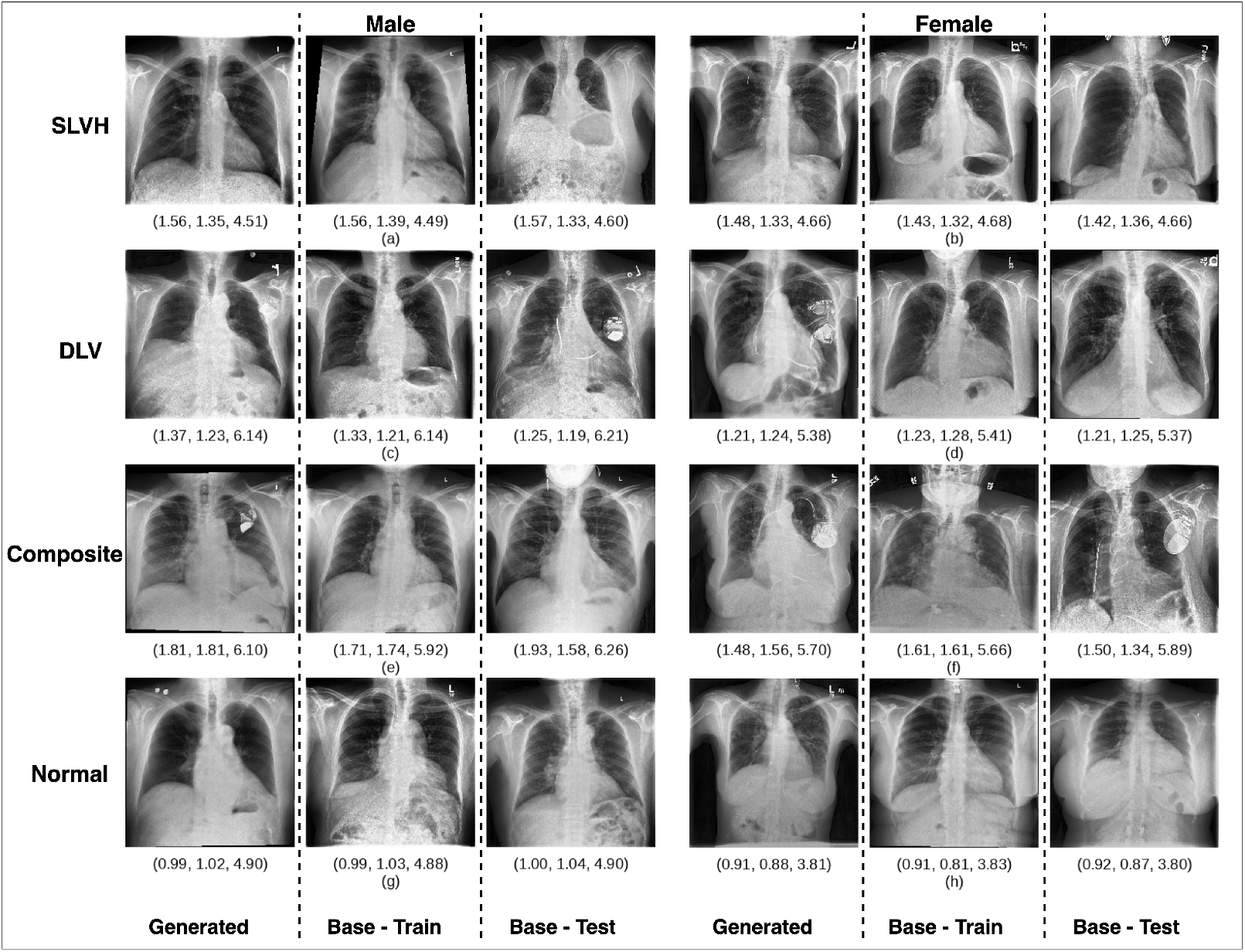
Visual comparison of generated images and real images using cross-matched features. Comparison of generated CXR images with cross-matched training and testing images based on age, sex, and echocardiogram measurements of IVSd, LVIDd, and LVPWd (displayed below each image). Each row of three images corresponds to a different grouping based on label (DLV, SLVH, composite, and none) and sex (male, female). The generated images can be observed to resemble the real images, demonstrating the model’s ability to produce realistic and demographically accurate images. A more detailed comparison of these images can be found in Supplementary Table 3.

### 2.4 Baseline Diagnostic Inference Results

We first established a baseline diagnostic performance by training a CNN model on the base dataset. The model produced an area under the receiver operating characteristic curve (AUROC) (Figure 3A) of 0.73 (95% CI of 0.71-0.76) for SLVH, 0.82 (95% CI of 0.79-0.84) for DLV, and 0.78 (95% CI of 0.76-0.80) for composite. The model produced an area under the precision-recall curve (AUPRC) of .45 (95% CI of .41-.48) for composite (Figure 3B). The full set of performance metrics can be observed in Supplementary Table 2.

**Fig. 3.**
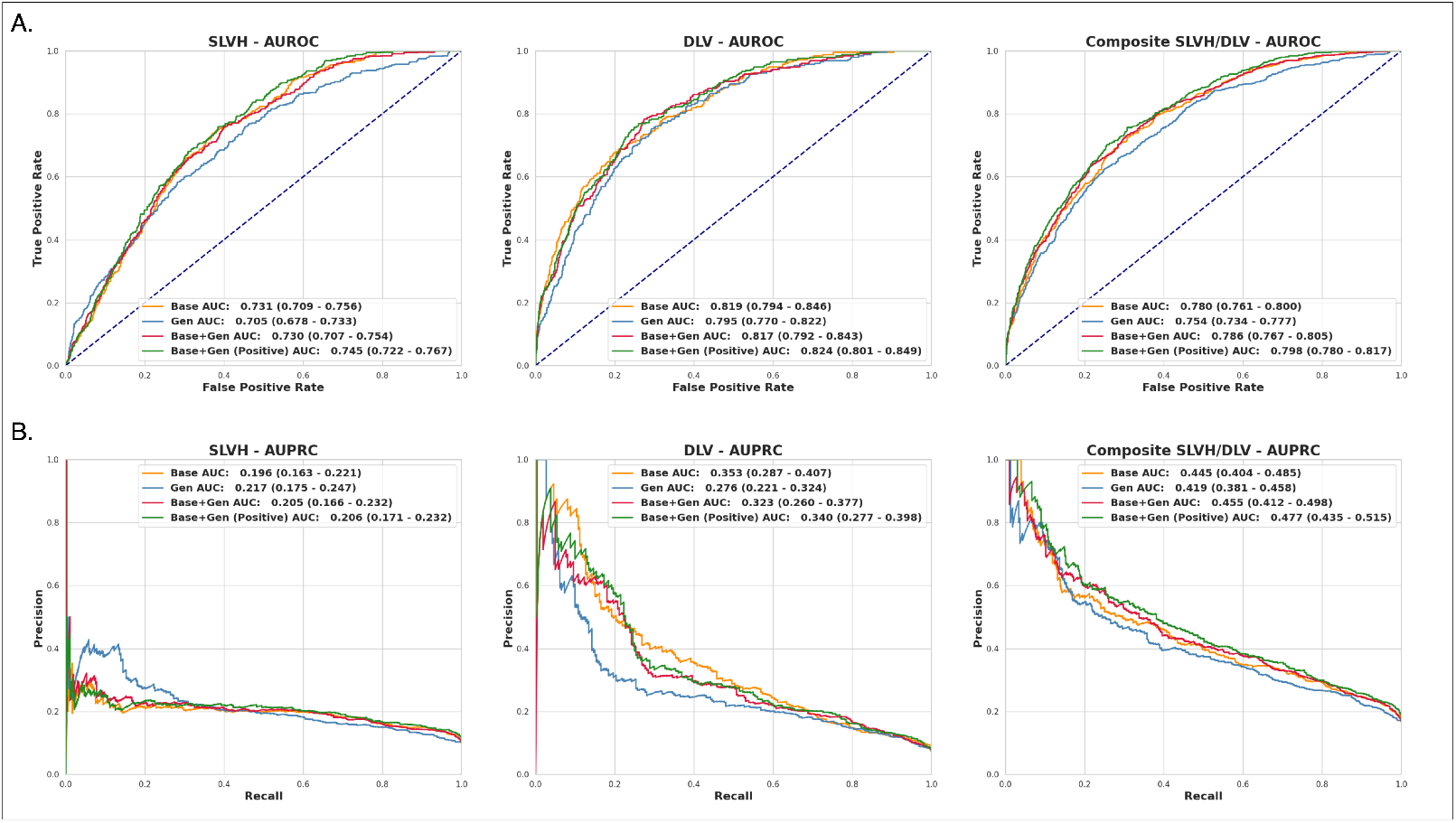
AUROC and AUPRC for the labels of SLVH, DLV, and composite across the four diagnostic models for the CheXchoNet dataset. **a)** The ROC curves and corresponding AUROC metrics for each of the diagnostic models for labels SLVH (left), DLV (middle), and composite (right). Each plot shows the performance of the models trained on the base (Base), generative (Gen), combined base and generative (Base+Gen), and combined base and positive generative samples (Base+Gen(Pos)). The Base+Gen(Pos) model produced the best overall performance with an AUROC of 0.80 (95% CI of 0.78-0.82, 2.3% above the base model) for the composite label. **b)** The precision-recall curves and corresponding AUPRC metrics for each of the diagnostic models for labels SLVH (left), DLV (middle), and composite (right). Each plot shows curves for each of the models listed above. The Base+Gen(Pos) model had the best overall performance with an AUPRC of .48 (95% CI .44-.53, 7.2% above baseline) for the composite label.

### 2.5 Generative Diagnostic Inference Results

Next, we assessed the diffusion model’s ability to encode demographic and echocardiogram data within generated images by training a model on this synthetic dataset. The model produced an AUROC of 0.71 (95% CI of 0.68-0.74, 3.6% below the base model) for SLVH, 0.80 (95% CI of 0.77-0.83, 2.9% below the base model) for DLV, and 0.75 (95% CI of 0.74-0.77, 3.3% below the base model) for composite (Figure 3A). The model produced an AUPRC of .42 (95% CI of .38-.46, 5.8% below the base model) for composite (Figure 3B). The results showed that the performance of the synthetic dataset was comparable to the base dataset, indicating that the diffusion model effectively encoded the input features.

### 2.6 Combined Diagnostic Inference Results

We evaluated two different combinations of synthetic and real data. The first combined the base data with all samples from the generated data, while the second combined the base data with only the positive samples from the generated data. The datasets were resampled with replacement such that they contained the same number of samples1. Using the total combined dataset, the model produced an AUROC of 0.73 (95% CI of 0.71-0.76, .15% below the base model) for SLVH, 0.82 (95% CI of 0.79-0.84, .3% below the base model) for DLV, and 0.79 (95% CI of 0.77-0.81, 0.7% above the base model) for composite (Figure 3A). The model produced an AUPRC of .46 (95% CI .42-.49, 2.2% above baseline) for composite (Figure 3B).

Using the focused combined dataset containing only positive generated samples, the model produced an AUROC of 0.75 (95% CI of 0.72-0.77, 1.8% above the base model) for SLVH, 0.82 (95% CI of 0.79-0.84, .5% above the base model) for DLV, and 0.80 (95% CI of 0.78-0.82, 2.3% above the base model) for composite (Figure 3A). The model produced an AUPRC of .48 (95% CI .44-.53, 7.2% above baseline) for composite (Figure 3B). The results showed that diagnostic performance can be increased through the selective sampling of synthetic data.

### 2.7 Internal Cohort Analysis

The performance of the models was validated on an internally collected dataset of patients from the University of Pennsylvania Health System. The internal cohort consisted of 315 CXRs from 265 patients and followed a similar age distribution to the CheXchoNet evaluation dataset, although was more evenly distributed across gender, as can be seen in Table 1. The dataset was constructed to have a more even distribution of labels, with 60% of samples having a positive composite label and 40% having no label. Each of the four diagnostic models was evaluated on this dataset. Overall, the models performed better on this dataset compared to the baseline CheXchoNet dataset and followed a similar trend with the base and focused combined models producing the best performance as can be seen in Figure 4. Both models producing an AUROC of 0.84 (95% CI of 0.80-0.89) on the composite label while the base model produced a better overall AUPRC of 0.87 (95% CI of 0.82-0.93) compared to 0.86 (95% CI 0.80-0.91) for the focused combined model.

**Table 1.**
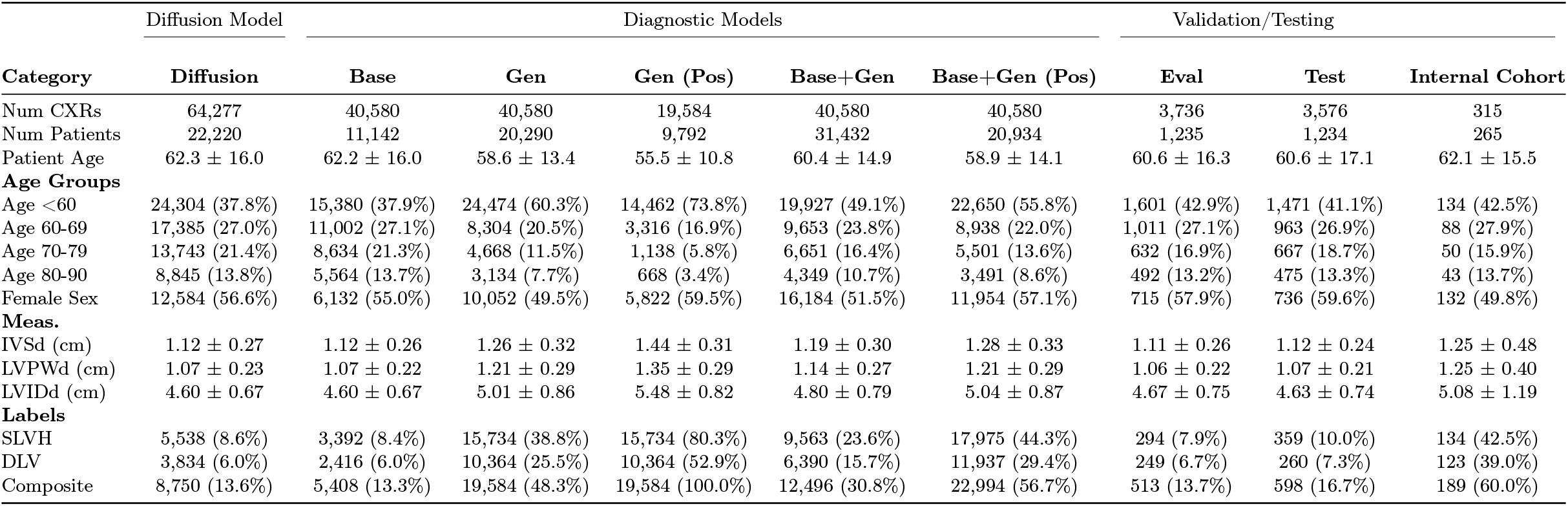
Dataset metrics for the base, generated, combined, testing and evaluation datasets. The Diffusion dataset was created by randomly sampling 90% of patients within the complete CheXchoNet dataset. The diagnostic model were trained on the Base, Gen, Base+Gen, and Base+Gen(Pos) datasets. The Base dataset was created by randomly sampling 20,290 images and corresponding features from the Diffusion dataset. This number was selected to match the number of unique samples generated by the diffusion model for the Gen dataset. All of the datasets were sampled with replacement to produce the final total size of 40,580 samples, to match the size of the Base+Gen dataset. The Gen (Pos) was not used for training a diagnostic model, it was instead combined with the Base dataset to produce the Base+Gen(Pos) dataset which was used to train a diagnostic model. The Eval dataset was used for model validation and the Test dataset was used to compute performance metrics.

**Fig. 4.**
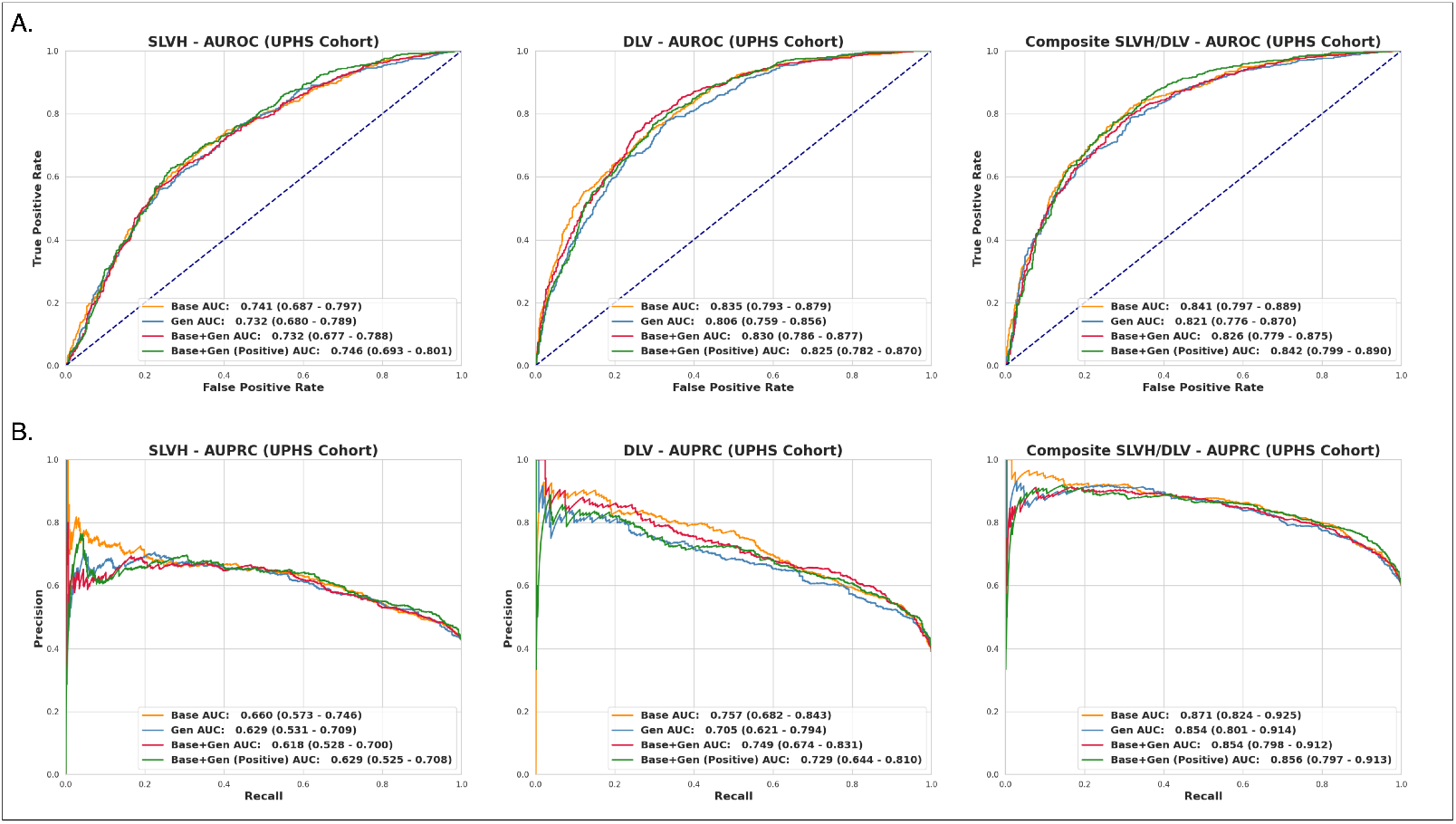
AUROC and AUPRC for the labels of SLVH, DLV, and composite across the four diagnostic models for the internal cohort collected at UPHS. **a)** The ROC curves and corresponding AUROC metrics for each of the diagnostic models. The models achieve similar performance, with Base and Base+Gen (Pos) producing the best overall results. **b)** The precision-recall curves and corresponding AUPRC metrics for each of the diagnostic models. The Base model produces the best overall results.

### 2.8 Subgroup Analysis

The performance of the models was further evaluated across different age subgroups to assess the impact of sampling the generated dataset for increased representation of patients under the age of 60. As displayed in Figure 5, models trained on the combined real and generated data outperformed the base model in all subgroups except for patients aged 80-90. The largest performance improvement was observed in patients under the age of 60. Specifically, the model trained on a combination of real data and positive generated samples achieved an AUROC of 0.86 (95% CI .84-.88), reflecting a 3.7% improvement over the base model. Improvements in other age subgroups were more modest, with increases of 1.5% and 1.4% for the 60-69 and 70-79 age groups, respectively, while the 80-90 age group saw a 1.5% decrease in performance.

**Fig. 5.**
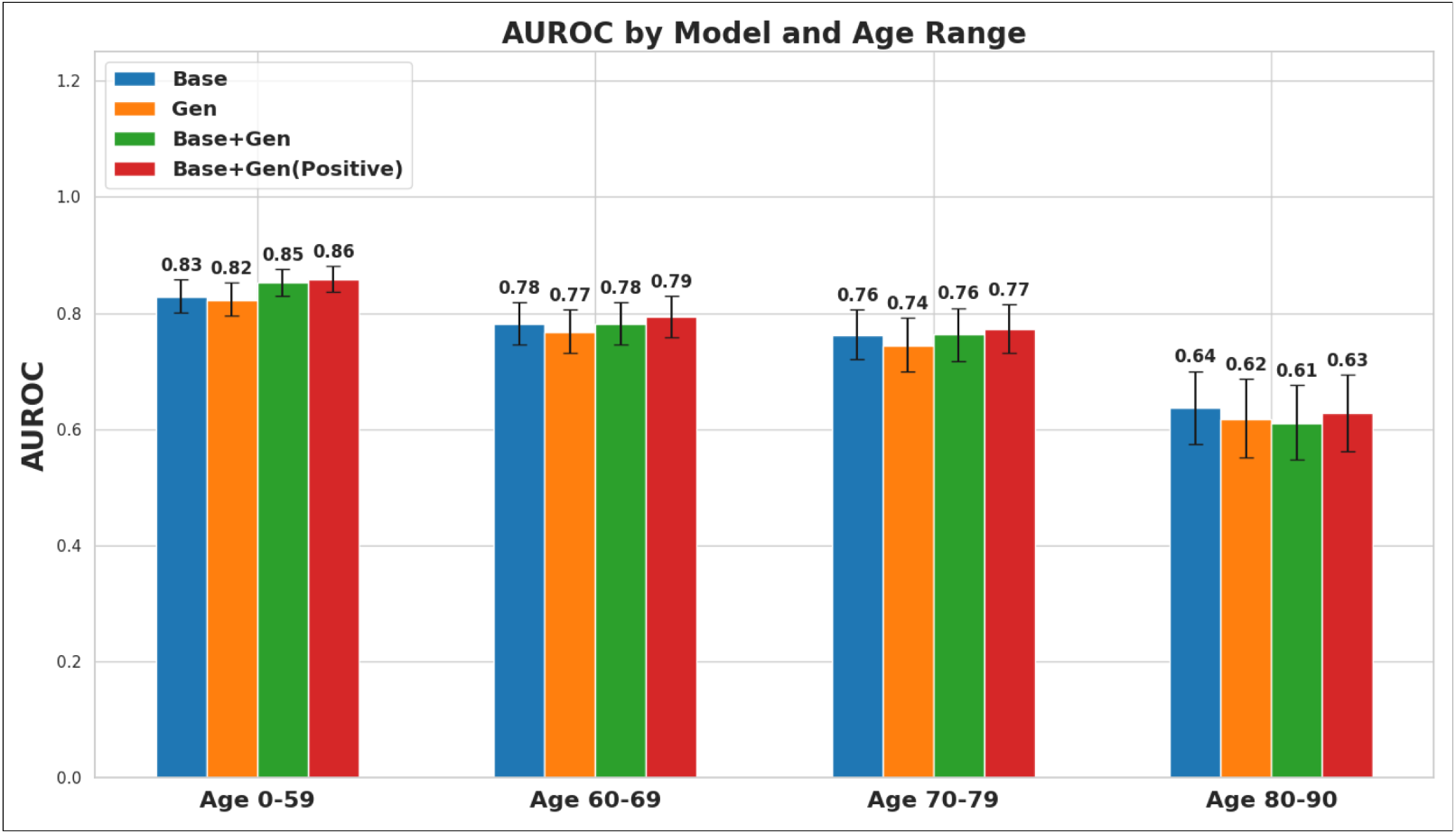
AUROC comparisons of the composite label across different age groups for each of the diagnostic models. Comparison of AUROC metrics for each of the four diagnostic models stratified by age group. The Base+Gen (Positive) model consistently outperforms the other models across all age groups besides 80-90 years. The model shows the most significant improvement in the under 60 age group with an AUROC of 0.86 indicating better discrimination within this subgroup as compared to the other groups.

**Fig. 6.**
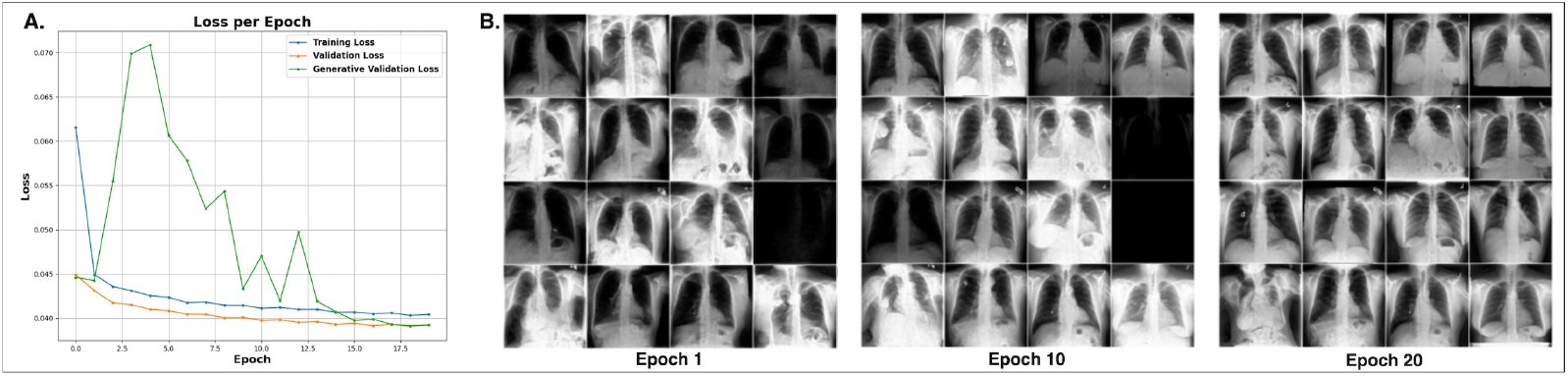
Training loss plots and evolution of generated images across epochs. **a)** Depicts the training progression of the diffusion model, showing the training loss, validation loss, and generative validation loss. The training and validation losses are computed using perceptual loss, while the generative validation loss is calculated by generating a batch of images from the validation dataset and computing the MSE loss between the generated and original images. All three loss values converge and stabilize around epochs 15-20, indicating a steady state in training. **b)** Shows the progression of generated validation images across different epochs, with image quality stabilizing at a high level between epochs 15-20.

**Fig. 7.**
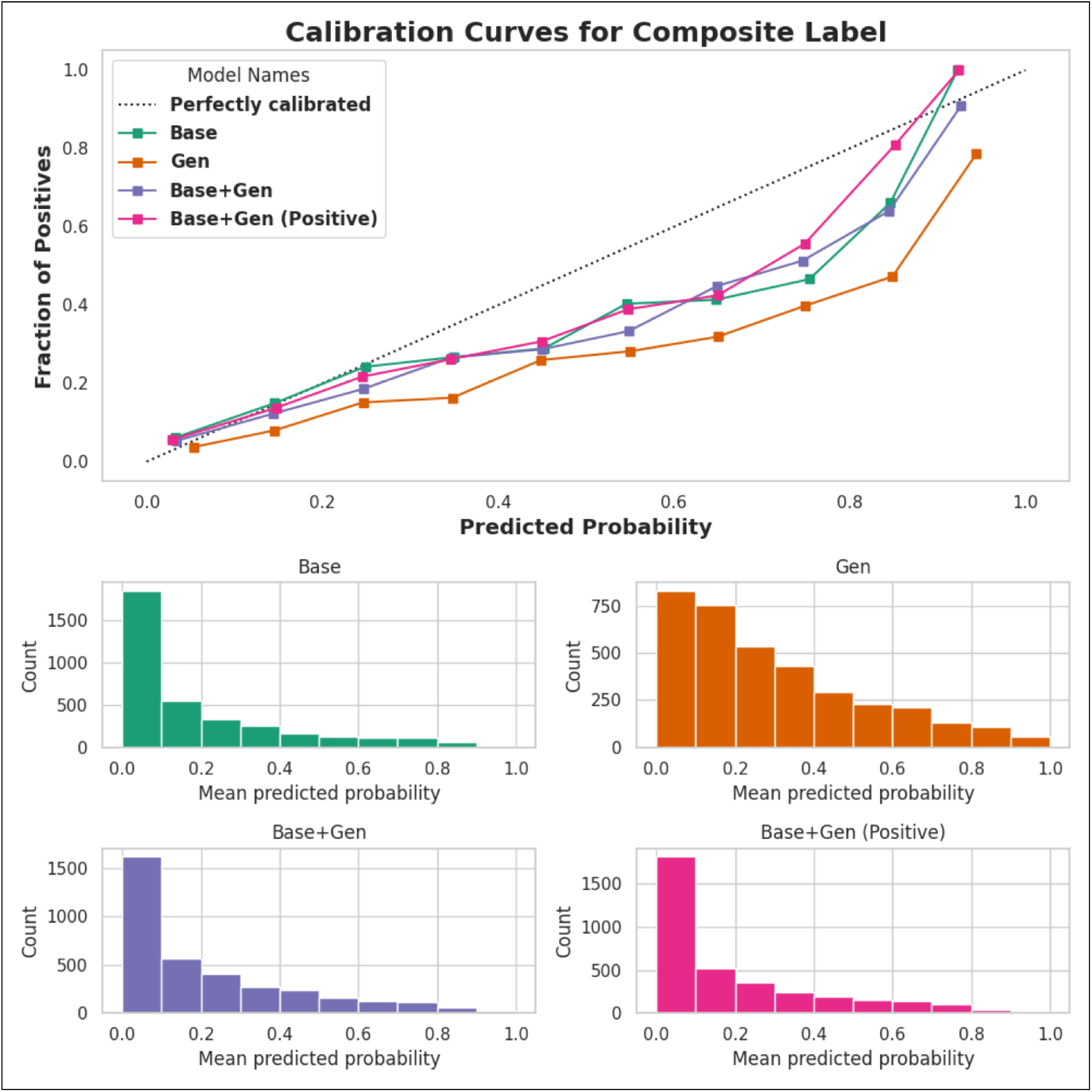
Calibration curves for the composite label across the four diagnostic models. The calibration plot compares the predicted probabilities of positive cases for the composite label against the fraction of positive results, with the dotted line representing a perfectly calibrated model. The graph shows calibration results for the four different models of Base, Gen, Base+Gen, and Base+Gen (Positive). The Base+Gen (Positive) model demonstrates the closest alignment to the optimal calibration, indicating more confident predictions. In contrast, the Gen model shows underconfident predictions more focused in the higher probability ranges. The histogram plots show the distribution of predicted probabilities for each model.

**Fig. 8.**
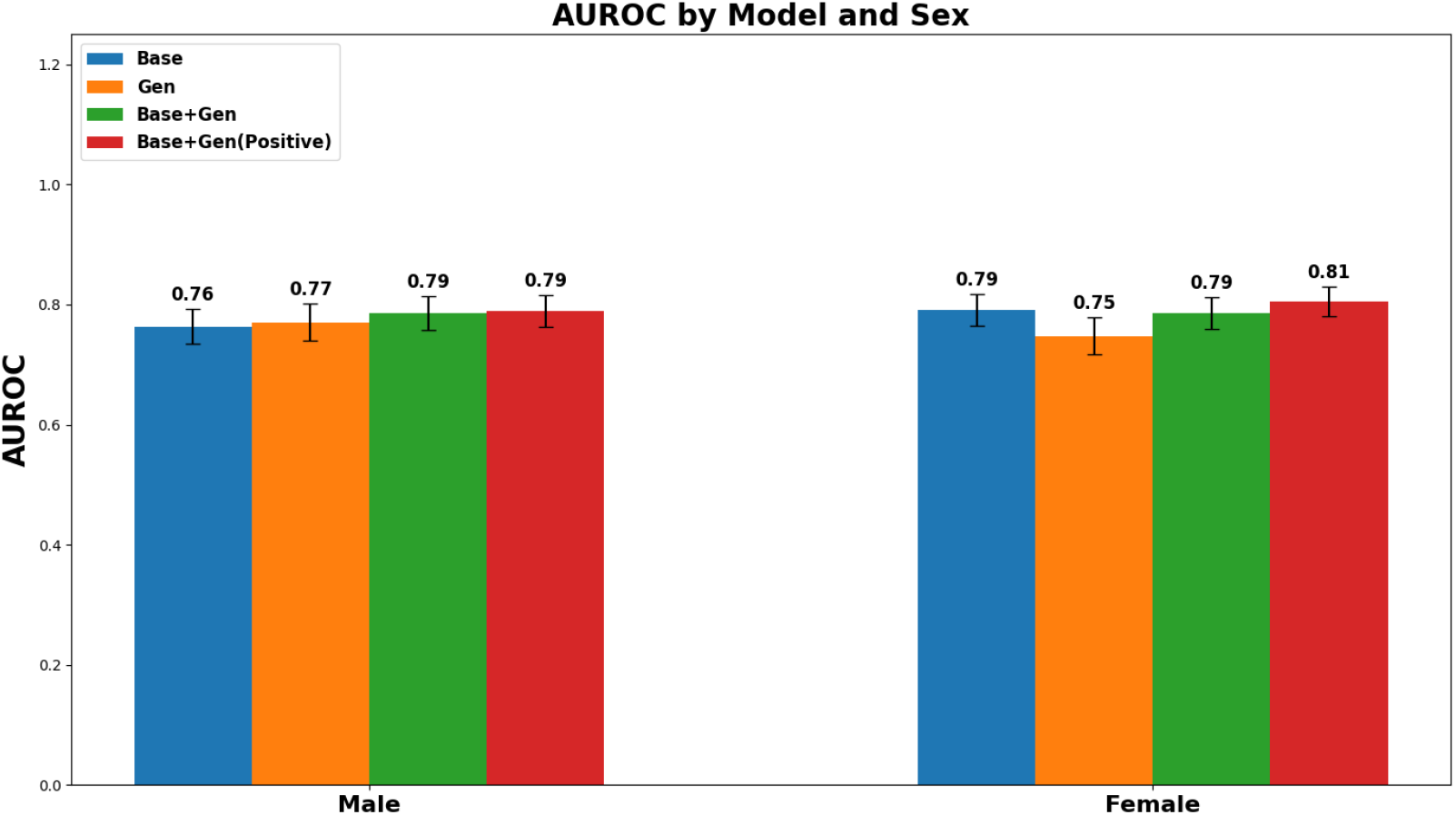
AUROC comparisons of the composite label stratified by sex for each of the diagnostic models. Comparison of AUROC metrics for each of the four diagnostic models stratified by sex. The Gen model shows improved performance for males compared to females; however, the Base+Gen model shows equal performance across both demographics. The Base+Gen (Positive) model consistently outperforms all other models for both males and females.

We additionally evaluated the diagnostic models across males and females (Supplementary Figure 8). We found that the model trained on generated data performed better on males than females; however, the model trained on the combined real and generated dataset produced similar performance between the two groups with a mean AUROC=0.79. The model trained on the combined real and positive generative samples displayed the best performance, achieving an AUROC=0.79 (95% CI of 0.76-0.82) on males and AUROC=0.81 (95% CI of 0.78-0.83) on females.

## 3 Discussion

In this work, we developed a diffusion model that encodes the joint distribution of demographic and echocardiogram measurements, enabling the generation of realistic and diverse images. We used this model to create a synthetic dataset with a distinct distribution from the original dataset, specifically by increasing the proportion of patients under 60 and the prevalence of SLVH and DLV. Our results demonstrated that training a deep learning model exclusively on synthetic data can closely approximate the performance of models trained on real data. Moreover, combining synthetic and real data led to improved overall diagnostic accuracy, with the most significant gains observed in the targeted demographic. Finally, we found that focusing on positive samples within the synthetic dataset resulted in the most substantial performance improvements.

This study serves as an extension of the previous work of developing the CheXchoNet [50] dataset and deep learning methodologies to detect structural heart disease [39]. Our results show how CheXchoNet can be further leveraged to create a generative model that has broad applicability. Most directly, this work further validates the use of diffusion models for creating synthetic datasets as has been demonstrated by previous studies [7, 8, 10, 21, 24–26]. This work distinguishes itself from previous studies by the incorporation of both categorical and continuous variables into the model, and leveraging these distributions to increase the signal of positive samples across a targeted demographic.

Before analyzing the downstream benefits of the model, we first assessed the quality of images within the synthetic dataset. Our analysis showed that the diffusion model was capable of producing both high-quality and diverse images. The synthetic dataset’s FID score of 9.62, compared to the training dataset, indicated that the generated images are similar in appearance to the base distribution and that the model is capable of producing high-quality images. Additionally, the synthetic dataset’s FID score of 13.5, compared to the testing dataset, suggests a slight reduction in quality and some overfitting to the training data. Despite this, the strong FID scores demonstrate the model’s overall ability to capture the distribution of CXRs and produce realistic images. Finally, the similar IS metrics across the synthetic, training, and testing datasets confirm that the diversity of the generated images matches that of the real data.

Our analysis of the diagnostic models trained across the base, synthetic, and combined datasets showed robust performance with the synthetic data. The diagnostic model trained on the synthetic dataset alone performed similarly to the base model, with results differing by only a few percentage points, indicating that synthetic data can approximate real data effectively. The diagnostic model trained on the combined dataset showed slight improvements in AUROC and AUPRC, compared to the base model. However, the base dataset combined with positive samples from the generated dataset showed substantial improvement in nearly all measured metrics. Notably, the diagnostic model trained on this dataset had increases in AUROC and AUPRC of 2.3% and 7.2%, respectively. This effect was most pronounced in patients under the age of 60, which showed an increase in AUROC of 3.7%. Additionally, the model trained on the combined datasets demonstrated similar performance between males and females, with AUROCs of 0.79 and 0.81, respectively. These results indicate that synthetic data can improve diagnostic model performance and that targeted synthetic datasets can be effectively designed to increase accuracy across specific populations or demographics.

We validated the performance of the diagnostic models on an internal dataset to assess reproducibility. All models demonstrated improved performance on this dataset, with the most notable gains observed for the composite label. However, the combined synthetic diagnostic models produced similar or worse performance compared to the base model. This lack of improvement with synthetic data could be attributed to the older average age of the internal cohort, which would affect model generalizability, or be caused by an underpowered study. Despite this, the synthetic models maintained strong performance, supporting the reproducibility and usefulness of this approach for diagnostic tasks.

These results demonstrate that generative models can be a powerful tool for combatting data scarcity within medicine. While collecting diverse real-world data remains the most effective way to improve model performance [24], this study highlights how diffusion models can create datasets with focused distributions, allowing for models to be fine-tuned for specific patient populations. We additionally believe that diffusion models and synthetic data have broad applications in medicine beyond just improving diagnostic model performance. Privacy concerns often limit data sharing between hospitals and health systems, but generative models can produce high-quality synthetic datasets that facilitate sharing while preserving patient privacy [23, 26]. However, further research is needed to fully understand the differential privacy aspects of diffusion models, as there is evidence that they may inadvertently memorize training data [53].

Diffusion models have additionally shown significant potential in advancing patient care by enabling the reconstruction of imaging data from specific input features. While existing research has focused on reconstructing MRI and CT images [18, 54–57], our work shows that patient-specific data can be used to generate realistic CXRs. By conditioning the model on values such as age, sex, and echocardiogram measurements, we were able to produce images that were specific to and representative of individual patients. Our approach demonstrates that, with a robust and descriptive set of input features, diffusion models can generate realistic and clinically relevant images tailored to the unique profiles of patients.

While diffusion models show significant promise for clinical applications, their deployment has limitations. Generating high-quality data using the DDPM methodology is computationally intensive and requires highperformance graphics processing units (GPUs). For example, generating a batch of synthetic data requires 1,000 forward passes using the model. Alternative methodologies, such as denoising diffusion implicit models (DDIMs) [58], offer a more efficient alternative approach to generating images and warrants further investigation to assess the quality trade-offs between these methodologies. Although we used standardized metrics to evaluate image quality and diversity, we did not have radiologists review the generated images to assess their realism as this was beyond the scope of our paper which focused on demonstrating improved performance of a diagnostic deep learning model.

Looking forward, this work not only demonstrates the feasibility of enhancing diagnostic performance but also lays the groundwork for validating this approach on external datasets. There is significant potential to refine this methodology by developing diffusion models that encode a wider range of input signals, enabling the generation of outputs that more accurately reflect patient demographics and clinical conditions. Incorporating a more complex feature vector that includes additional demographic details, vital signs, and other diagnostic results such as electrocardiograms could significantly improve the fidelity and utility of generated data that more accurately reflects individual patients.

Overall, this work demonstrates the potential of diffusion models to generate realistic, diverse, and clinically relevant images, thereby addressing data scarcity and enhancing diagnostic model performance across targeted demographics.

## 4 Methods

### 4.1 Dataset Preparation

We used the CheXchoNet [50] dataset as the basis of our model training. The total dataset consisted of 71,589 CXRs collected from 24,689 different patients at the Columbia University Irving Medical Center (CUIMC) between January 2013 to August 2018. Each CXR was paired with a TTE performed within 12 months of each other. Inclusion criteria required each CXR to have at least one TTE pairing within the specified time frame. The dataset consisted of only posteroanterior (PA) films and were extracted in their complete DICOM format. Each DICOM was preprocessed by first cropping the image to a 1:1 aspect ratio then downsampling the image to 224×224 pixels using bicubic interpolation. The echocardiograms were accessed using the Syngo Dynamics system and the measures of IVSd, LVIDd, and LVPWd were extracted from the parasternal long axis view. Binary diagnosis labels for SLVH and DLV were determined using echocardiographic thresholds based on current guidelines. SLVH was defined as IVSd or LVPWd > 1.5 cm in men and > 1.4 cm in women, while DLV was defined as LVIDd > 5.9 cm in men and > 5.3 cm in women. A composite label indicated the presence of either condition [39, 50].

We initially divided the dataset into partitions for training, validation, and testing with partition sizes of 90%, 5%, and 5% randomized by patient to ensure no data leakage between testing and evaluation. The corresponding datasets consisted of 64,277 CXRs across 22,220 patients for training, 3,736 CXRs across 1,235 patients for validation, and 3,576 CXRs across 1,234 patients for testing. The full set of metrics for each of these partitions can be found in Table 2.

**Table 2.** Diagnostic model performance across the SLVH, DLV, and composite labels. Performance metrics for each of the four diagnostic models stratified by label, comparing brier loss, log loss, AUROC, AUPRC, precision, and specificity. Precision and specificity were both computed at a 50% recall threshold.

|  | Brier loss | Log loss | AUROC | AUPRC | Precision | Specificity |
| --- | --- | --- | --- | --- | --- | --- |
| **SLVH** |  |  |  |  |  |  |
| Base | 0.106 [0.098 - 0.113] | 0.343 [0.324 - 0.363] | 0.731 [0.706 - 0.760] | 0.196 [0.159 - 0.223] | 19.8% | 77.4% |
| Gen | 0.115 [0.110 - 0.120] | 0.375 [0.360 - 0.393] | 0.705 [0.681 - 0.735] | 0.217 [0.185 - 0.242] | 19.4% | 76.8% |
| Base+Gen | 0.106 [0.098 - 0.112] | 0.341 [0.325 - 0.362] | 0.730 [0.709 - 0.760] | 0.205 [0.178 - 0.233] | 20.4% | 78.2% |
| Base+Gen (Positive) | 0.103 [0.098 - 0.109] | 0.331 [0.306 - 0.350] | 0.745 [0.722 - 0.769] | 0.206 [0.172 - 0.234] | 21.4% | 79.5% |
| **DLV** |  |  |  |  |  |  |
| Base | 0.057 [0.052 - 0.064] | 0.209 [0.191 - 0.227] | 0.819 [0.794 - 0.841] | 0.353 [0.286 - 0.400] | 28.8% | 90.3% |
| Gen | 0.070 [0.066 - 0.076] | 0.244 [0.228 - 0.256] | 0.795 [0.770 - 0.825] | 0.276 [0.210 - 0.334] | 22.0% | 86.1% |
| Base+Gen | 0.058 [0.053 - 0.065] | 0.212 [0.193 - 0.229] | 0.817 [0.793 - 0.842] | 0.323 [0.260 - 0.371] | 27.5% | 89.7% |
| Base+Gen (Positive) | 0.058 [0.051 - 0.064] | 0.213 [0.190 - 0.231] | 0.824 [0.801 - 0.847] | 0.340 [0.253 - 0.391] | 27.8% | 89.8% |
| **Composite** |  |  |  |  |  |  |
| Base | 0.125 [0.117 - 0.134] | 0.396 [0.380 - 0.414] | 0.780 [0.759 - 0.797] | 0.445 [0.408 - 0.483] | 39.7% | 84.7% |
| Gen | 0.150 [0.142 - 0.155] | 0.461 [0.442 - 0.475] | 0.754 [0.735 - 0.773] | 0.419 [0.382 - 0.457] | 37.3% | 83.1% |
| Base+Gen | 0.124 [0.118 - 0.131] | 0.393 [0.369 - 0.411] | 0.786 [0.768 - 0.806] | 0.455 [0.418 - 0.491] | 40.7% | 85.4% |
| Base+Gen (Positive) | 0.119 [0.110 - 0.126] | 0.379 [0.357 - 0.405] | 0.798 [0.779 - 0.818] | 0.477 [0.441 - 0.527] | 43.1% | 86.7% |

**Table 3.** Complete demographic and inference results for cross-matched images. This table provides more detail for the images observed in Figure 2 including demographic data and inference results for each of the four diagnostic models for the provided label. The inference results show the mean-average probabilities generated by the diagnostic models for the Base, Gen, Base+Gen, and Base+Gen(Pos) datasets. While the Gen model tends to produce overly optimistic probabilities, the combined dataset models produce more balanced probabilities, effectively integrating characteristics of both the Gen and Base models.

| Demographics |  |  | Measurements |  |  | Label and Inferences |  |  |  |  |
| --- | --- | --- | --- | --- | --- | --- | --- | --- | --- | --- |
| Image | Age | Sex | IVSd | LVPWd | LVIDd | Ground Truth | Base | Gen | Base+Gen | Base+Gen(Pos) |
| A | 52 | M | 1.57 | 1.33 | 4.60 | SLVH | 2.2% | 39.8% | 7.6% | 7.9% |
| B | 68 | F | 1.42 | 1.36 | 4.66 | SLVH | 0.6% | 24.1% | 6.4% | 5.0% |
| C | 66 | M | 1.25 | 1.19 | 6.21 | DLV | 31.9% | 34.0% | 19.5% | 16.8% |
| D | 76 | F | 1.21 | 1.25 | 5.37 | DLV | 2.7% | 3.8% | 0.2% | 0.2% |
| E | 47 | M | 1.93 | 1.58 | 6.26 | Composite | 73.3% | 77.9% | 56.0% | 51.7% |
| F | 79 | F | 1.50 | 1.34 | 5.89 | Composite | 46.5% | 82.0% | 58.4% | 50.9% |
| G | 82 | M | 1.00 | 1.04 | 4.90 | Normal | 60.5% | 90.5% | 76.9% | 75.7% |
| H | 51 | F | 0.92 | 0.87 | 3.80 | Normal | 98.6% | 78.6% | 95.1% | 98.1% |

**Table 4.** Diagnostic model performance for the composite label across different age groups. Performance metrics for each of the four diagnostic models on the composite label stratified by age group. Reported metrics include comparing brier loss, log loss, AUROC, and AUPRC.

|  | Brier loss | Log loss | AUROC | AUPRC |
| --- | --- | --- | --- | --- |
| **Age < 60** |  |  |  |  |
| Base | 0.170 [0.147 - 0.190] | 0.527 [0.460 - 0.587] | 0.637 [0.574 - 0.700] | 0.285 [0.193 - 0.356] |
| Gen | 0.162 [0.141 - 0.181] | 0.510 [0.450 - 0.564] | 0.618 [0.552 - 0.686] | 0.281 [0.180 - 0.349] |
| Base+Gen | 0.162 [0.139 - 0.184] | 0.517 [0.445 - 0.582] | 0.610 [0.547 - 0.676] | 0.259 [0.164 - 0.322] |
| Base+Gen (Positive) | 0.161 [0.138 - 0.181] | 0.508 [0.443 - 0.571] | 0.628 [0.562 - 0.693] | 0.286 [0.192 - 0.356] |
| **Age 60-69** |  |  |  |  |
| Base | 0.137 [0.121 - 0.152] | 0.451 [0.401 - 0.497] | 0.782 [0.746 - 0.819] | 0.569 [0.498 - 0.634] |
| Gen | 0.157 [0.143 - 0.170] | 0.484 [0.450 - 0.518] | 0.767 [0.730 - 0.806] | 0.536 [0.467 - 0.604] |
| Base+Gen | 0.138 [0.122 - 0.152] | 0.447 [0.401 - 0.492] | 0.781 [0.745 - 0.819] | 0.586 [0.519 - 0.656] |
| Base+Gen (Positive) | 0.132 [0.117 - 0.146] | 0.438 [0.386 - 0.486] | 0.793 [0.758 - 0.830] | 0.610 [0.548 - 0.674] |
| **Age 70-79** |  |  |  |  |
| Base | 0.137 [0.120 - 0.153] | 0.424 [0.373 - 0.472] | 0.762 [0.721 - 0.806] | 0.419 [0.324 - 0.507] |
| Gen | 0.143 [0.128 - 0.157] | 0.442 [0.402 - 0.478] | 0.744 [0.700 - 0.792] | 0.431 [0.341 - 0.519] |
| Base+Gen | 0.131 [0.115 - 0.146] | 0.413 [0.367 - 0.455] | 0.764 [0.718 - 0.809] | 0.433 [0.347 - 0.513] |
| Base+Gen (Positive) | 0.131 [0.114 - 0.148] | 0.409 [0.365 - 0.453] | 0.773 [0.731 - 0.815] | 0.435 [0.345 - 0.518] |
| **Age 80-90** |  |  |  |  |
| Base | 0.170 [0.147 - 0.190] | 0.527 [0.460 - 0.587] | 0.637 [0.574 - 0.700] | 0.285 [0.193 - 0.356] |
| Gen | 0.162 [0.141 - 0.181] | 0.510 [0.450 - 0.564] | 0.618 [0.552 - 0.686] | 0.281 [0.180 - 0.349] |
| Base+Gen | 0.162 [0.139 - 0.184] | 0.517 [0.445 - 0.582] | 0.610 [0.547 - 0.676] | 0.259 [0.164 - 0.322] |
| Base+Gen (Positive) | 0.161 [0.138 - 0.181] | 0.508 [0.443 - 0.571] | 0.628 [0.562 - 0.693] | 0.286 [0.192 - 0.356] |

We further validated the performance of the models across an internally collected dataset consisting of UPHS patients. This dataset was collected by first identifying all patients who had a CXR performed within 12 months of a TTE between September 1st, 2023 and September 1st, 2024. A total of 312,610 possible CXR studies were identified. Labels of SLVH, DLV, and composite were assigned to each of these studies using the matching TTE measurements and thresholds specified by CheXchoNet. We then randomly sampled 200 normal CXRs, 100 with strictly LVH, 100 with strictly SLVH, and 100 with both a composite of DLV and SLVH for a total of 500 studies. Out of those studies, 185 were not able to be included because of privacy reasons or difficulty accessing the image, leaving a total of 315 studies which were included in our analysis. The full set of metrics for this cohort can be found in Table 1.

### 4.2 Diffusion Model Architecture and Training

The goal of the work was to develop a generative model capable of producing realistic images conditioned on the provided input data. We selected to use a DDPM [4, 5] because of its demonstrated ability to produce high quality outputs and the ease of training compared to other architectures such as GANs [59]. Training a diffusion model consists of an initial forward process where gaussian noise is sequentially added to an image followed by a reverse process where the model tries to estimate and incrementally remove the added noise with goal of recreating the original image [60]. We used a cosine-beta scheduler [5] to add noise to the images and set the maximum number of steps to be 1,000. We conditioned the model by providing an input feature vector containing the demographic and echocardiogram measurements which allowed the model to learn to generate images specific to the provided features. Sex was converted to a one-hot input vector while all continuous variables were normalized using a z-score taken from the training dataset.

Our selected model used a U-Net [61] architecture consisting of encoder and decoder components and has been commonly used for medical image segmentation [62]. The U-Net took as input a 224×224 pixel image with one grayscale channel and output a 224×224 vector representing the estimated noise added to the image.

Our model consists of multiple downsampling and upsampling blocks with residual connections and attention layers. The model additionally takes as input a timestep embedding which provides temporal context during the step-wise process and a state embedding which encodes the conditional features.

We trained the model using a combination of mean squared error (MSE) loss and perceptual loss [63, 64]. MSE loss is computed by taking the mean of the squared error between the noise estimated by the model and actual noise added to the image. Perceptual loss is computed by calculating differences between high-level features of the predicted and target images extracted using a pre-trained VGG16 [65] model. The final loss is computed using a weighted average between MSE loss and perceptual loss. This loss function was used to ensure that generated images were both accurate at the pixel level and perceptually similar to the original images. This method has been shown to improve overall image generation quality. We recorded this loss for both the training images during each gradient update step and for validation images during scheduled intervals. We additionally recorded a generative validation loss which measured the MSE between a fixed set of validation images and generated images. The generated images were conditioned using the corresponding features i.e. demographics, echocardiogram measures from the validation images.

We trained the model across 20 epochs with a batch size of 16 and used the AdamW optimization algorithm [66] to perform gradient descent. The models were trained using a single NVIDIA A100 GPU. Supplementary Figure 6 shows the loss per each epoch and the progression of generated validation images. As can be observed, the model’s loss converged around epoch 15 which corresponded with stabilization of image quality.

### 4.3 Synthetic Dataset Generation and Evaluation

The synthetic dataset was generated using the fully-trained diffusion model. The process began by sampling random noise from a standard normal distribution and conditional features from their respective distributions. These values were then input into the diffusion model, which conditionally denoised the images over 1,000 iterations, guided by the provided features and timestep embeddings. During each iteration, the model estimated the noise present in the image, which was subsequently removed by the scheduler, and the image was resampled from the resulting distribution. This iterative process continued for the specified number of steps to produce the final generated image [60].

We generated the input features by randomly sampling from normal distributions for the continuous variables (age, IVSd, LVIDd, LVPWd) and randomizing between males and females. For age, we sampled from a normal distribution with a lower mean than that of the training dataset, and higher means for IVSd, LVIDd, and LVPWd. Overall, we produced a synthetic dataset consisting of 20,290 unique CXRs with full features as listed in Table 1. We added binary labels using the same thresholds specified by CheXchoNet.

We evaluated the quality and diversity of the synthetic dataset using FID [67] and IS [68] metrics. FID compares the mean and covariance of feature vectors extracted from a pre-trained Inception network [52] between a synthetic and real dataset with a lower score indicating higher-quality images. We performed this comparison between the synthetic dataset and both the training and testing datasets. IS evaluates both diversity and quality by passing images through an inception network and computing the entropy of the predicted classes.

### 4.4 Diagnostic Model Training and Evaluation

We used the same model architecture and training process detailed by Bhave et al. [39] for our diagnostic evaluation. The goal of the diagnostic evaluation was to train a CNN to detect the presence of SLVH, DLV, and the composite label. The methodology used the DenseNet-121 architecture [69] to process the input image and produce a feature vector. The feature vector was combined with the patient’s demographic data which was then used to estimate the three continuous variables of LVIDd, IVSd and, LVPWd which were subsequently used to compute the probabilities of the binary labels [39].

We created four different datasets for this evaluation. The first dataset consisted of the base images, the second dataset consisted of the synthetic images, the third dataset consisted of the base and synthetic images, and the final dataset consisted of the base images combined with the positive samples from the generated dataset. As shown in Table 1, fewer base images were used for training the diagnostic model compared to the diffusion model. This discrepancy arose because we downsampled the base dataset to match the number of unique CXRs in the generated dataset in order to standardize model training. Since the synthetic dataset consisted of 20,290 CXRs, we used this as the sampling benchmark when selecting images from the real dataset. We then resampled with replacement all datasets to ensure they had the same total number images. Since the combined dataset contained 40,580 images (20,290 real and 20,290 synthetic), we resampled all other datasets to match this number.

We trained the diagnostic models using the same parameters with a batch size of 32 across 10 epochs using the Adam optimizer [70] with early stopping. The models were trained using a single NVIDIA L4 GPU. For each dataset, we performed a total of 5 training runs and stored the model from each run which produced the lowest validation loss, as measured on the validation dataset. For our final evaluation, we used the withheld testing dataset to compute probabilities for each class of SLVH, DLV, and composite. Final probabilities were computed by mean-averaging the outputs of the five models for each dataset. We compared the models using standard metrics including log loss, brier loss, AUROC, and AUPRC. We computed the specificity and precision for each of the models using a fixed recall of 50%. We used bootstrapping methods to compute confidence intervals for the metrics. We performed a similar analysis across the different subgroups of patient ages.

## 5 Data Availability

This study uses the CheXchoNet dataset which is hosted on Physionet under a restricted access use policy. The dataset is available to registered users who sign the specified data use agreement. The study also uses an internal dataset from the University of Pennsylvania Health System. This dataset cannot be shared for ethical/privacy reasons.

## 6 Code Availability

The full code used to develop the diffusion model and generate the synthetic dataset is available at: https://github.com/cstreiffer/cxr_ddpm. The code used to recreate the diagnostic experiment is available at: https://github.com/sbhave77/CheXchoNet.

## 7 Acknowledgments

The research for this work was supported in part by the National Institutes of Health grant no. R01HL171709, R01HL169378, and P41EB029460. M.G.L. received support from the Doris Duke Foundation (Award 2023-0224), and US Department of Veterans Affairs (IK2-BX006551). This work was supported internally by the Department of Radiology and Center for Cardiovascular Informatics.

## 8 Author Contributions

C.S. implemented the diffusion model, generated the synthetic data, and performed the diagnostic model evaluation across listed datasets. E.A. and W.W. helped to construct the internal dataset for further model validation. M.L., W.W., and E.A. provided feedback and guidance on the core components of the manuscript. All authors contributed to the drafting of the manuscript and approved the final version.

## 10 Supplementary Material

## Footnotes

1 Please see methods for a more detailed description of how the datasets were assembled.

